# Detection of Acute Myocardial Infarction Using Deep Learning on Lead-I ECG Data

**DOI:** 10.1101/2024.10.15.24315544

**Authors:** Parmida Davarmanesh, Qian Lin, Irene Tenison, Gabriel Jabbour, Ridwan Alam

## Abstract

Myocardial Infarction (MI) is a major global health concern due to its high mortality and morbidity rates. Early detection of MI is crucial for timely medical intervention and improved patient outcomes. In this study, we investigate the feasibility of predicting MI using lead-I of electrocardiogram (ECG) data, with a focus on practical applications for wearable monitoring systems. Utilizing the PTB-XL dataset, which includes a comprehensive collection of 12-lead ECG recordings with both normal and various MI samples, we employ deep learning techniques to develop a binary classification model. For MI detection using lead-I, we achieved an AUC of 0.92 and an AUPR of 0.82 on the test set. In comparison, using 6-lead and 12-lead configurations both resulted in an AUC of 0.99. These findings demonstrate the potential for detecting MI using only lead-I, as measured by wearable devices. This advancement could significantly enhance clinical outcomes for MI patients by enabling timely detection and intervention.

## I. Introduction

Myocardial infarction (MI), commonly known as a heart attack, is the leading cause of death globally, affecting more than 8.9 million individuals annually [1]. Timely diagnosis of MI is of utmost importance, because delays in diagnosing these events can significantly increase mortality rates. In patients with MI presenting to the emergency department (ED), every 30-minute delay in treatment increases the risk of oneyear mortality by approximately 7.5*%* [2]. While MI is often associated with symptoms such as chest pain or discomfort, strikingly, it is estimated that 70-80*%* of cases, known as silent MI, lack any symptoms, increasing the risk of mortality and other long-term adverse health outcomes due to delayed medical intervention. [3], [4].

Currently, when patients present with symptoms suggestive of an MI, the first-line diagnostic test is an electrocardiogram (ECG), which measures the altered electrical activity of the heart [5]. In a hospital setting, ECGs are measured by putting a number of electrodes on multiple fixed locations of the patients’ body, resulting in 12 leads (directions) that provide a comprehensive view of the heart’s electrical activity.

One way the detection of MI can be accelerated is through the use of wearable devices such as smartwatches, which have already shown to be effective in conditions such as detecting atrial fibrillation [6], [7]. Smartwatches primarily measure a single lead out of 12 standard leads, known as lead-I.With the motivation to transfer clinically developed solutions toward outpatient monitoring, researchers proposed lead-I ECG-based deep learning solutions for applications ranging from arrhythmia detection to drug-effect tracking [8]–[10]. Detecting MI using only lead-I, as opposed to relying on a 12-lead ECG typically available only in hospital settings, could potentially enable MI patients to receive medical intervention much sooner than they would otherwise, thereby improving outcomes significantly. Our goal in this work is to detect MI using only lead-I of ECG data. Figure 1 illustrates the clinical motivation behind our work.

**Fig. 1.**
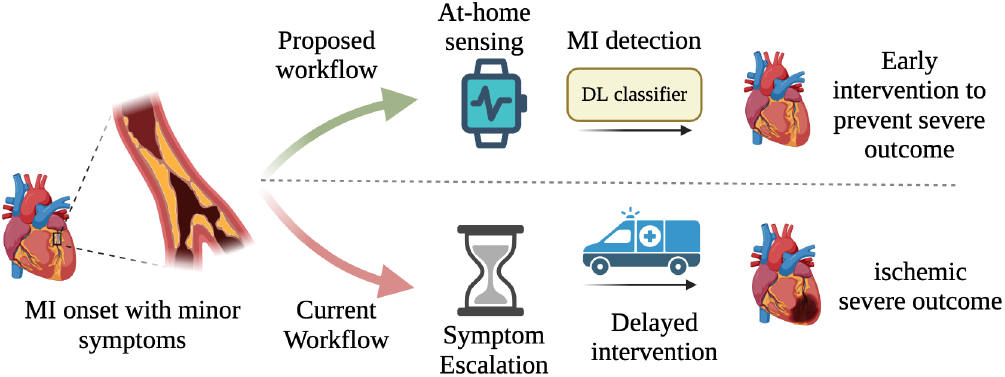
Clinical motivation and downstream application of our work

To our knowledge, there has been no prior study on the potential of lead-I alone in detecting MI. A number of works have investigated the task of MI detection using traditional Machine Learning (ML) methods. [11] used a K-nearest neighbor model on hand selected features from all 12 leads of ECG data, and achieved an accuracy of 99*%*. [12] showed that lead-I can be used to improve the assessment of the relative risk of MI by looking at certain features of the ECG signal using a proportional hazards regression model. Several studies have demonstrated superior performance of deep learning (DL) models compared to cardiologists in detecting MI from the 12-lead ECG readings [13]. Some works have also demonstrated the potential of detecting myocardial infarction (MI) from single-lead (other than lead-I) ECG using DL. For instance, using a single lead-V2 ECG with a 1-D convolutional neural network, MI detection achieved 90.5*%* accuracy [14]. Another study applied Gabor filters with an eight-layer CNN and weight balancing on lead-II ECG, reporting accuracy, specificity, and sensitivity values of 99.55*%*, 99.67*%*, and 99.27*%*, respectively, on the PTB dataset [15]. Similarly, a pretrained VGGNet model achieved an accuracy of 99.02*%*, sensitivity of 98.76*%*, and specificity of 99.17*%* [16]. Additionally, a 22-,layer CNN model reported accuracy, precision, F1 score, and recall of 98.84*%*, 98.31*%*, 97.92*%*, and 97.63*%*, respectively, utilizing focal loss to address data imbalance [17]. Despite these promising results, two main issues remain: reliance on manual feature extraction and limited generalizability due to small training datasets.

In this study, we utilize deep learning models trained on lead-I signals extracted from 12-lead ECGs to evaluate, as a proof of concept, whether lead-I alone contains sufficient information to detect MI, with remarkable clinical implications utilizing wearable monitoring devices.

## II. Methods

### A. Dataset

In this work, we utilized PTB-XL [18], a large publicly available dataset of 12-lead 10 second clinical ECG signals consisting of around 21000 records. Each record in this dataset has two hierarchical categories of superclass and subclass labels that are automatically extracted from clinical annotations. Since each record can have more than one diagnostic label, we only included the records with superclass labels “NORM” (negative class), and those with labels “MI” or “MI and STTC” as our positive class (STTC refers to “ST/T change” in ECG signals which is commonly seen within MI patients). MI patients with any other diagnostic class such as MI and ventricular hypertrophy (“HYP”) were excluded from our dataset. Additionally, in the PTB-XL dataset, each diagnostic label has a corresponding confidence score that is assigned based on the existence of certain words in the annotations. For example, the phrase “cannot be excluded” is assigned a confidence score of 15*%*, “probably” and “possible” get a score of 50*%*, and “consistent with” has a score of 100*%*. In order to ensure high label quality, we only included ECGs that are validated by at least one cardiologist and have 100*%* confidence scores. These exclusion criteria yielded a dataset of 5848 normal and 738 MI labeled ECGs which we used to perform binary classification. Figure 2 summarises the exclusion criteria and the patient demographics. Lastly, among the MI records, we used the “infarction stadium” variable to extract the chronicity of the MI: acute vs chronic MI, which often have different ECG presentations. Of our 738 MI records, we had 51 acute MIs, 369 chronic MIs, and the rest had unknown chronicity (could be either acute or chronic).

**Fig. 2.**
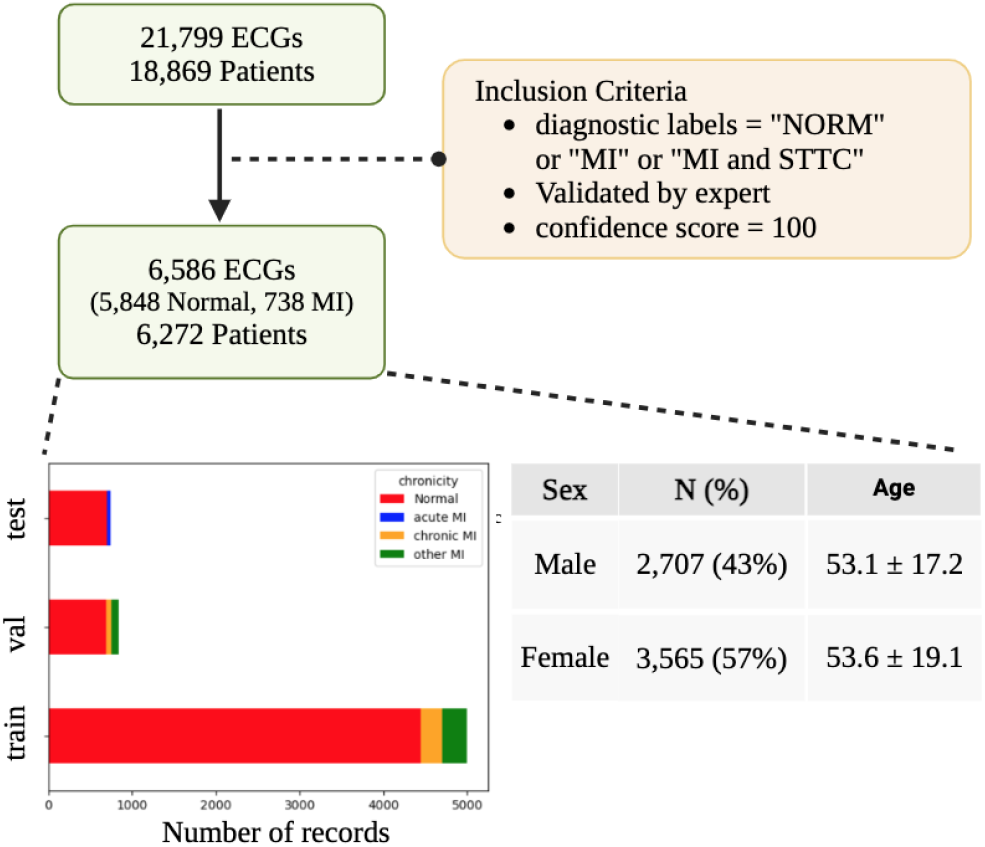
Exclusion Criteria

**Fig. 3.**
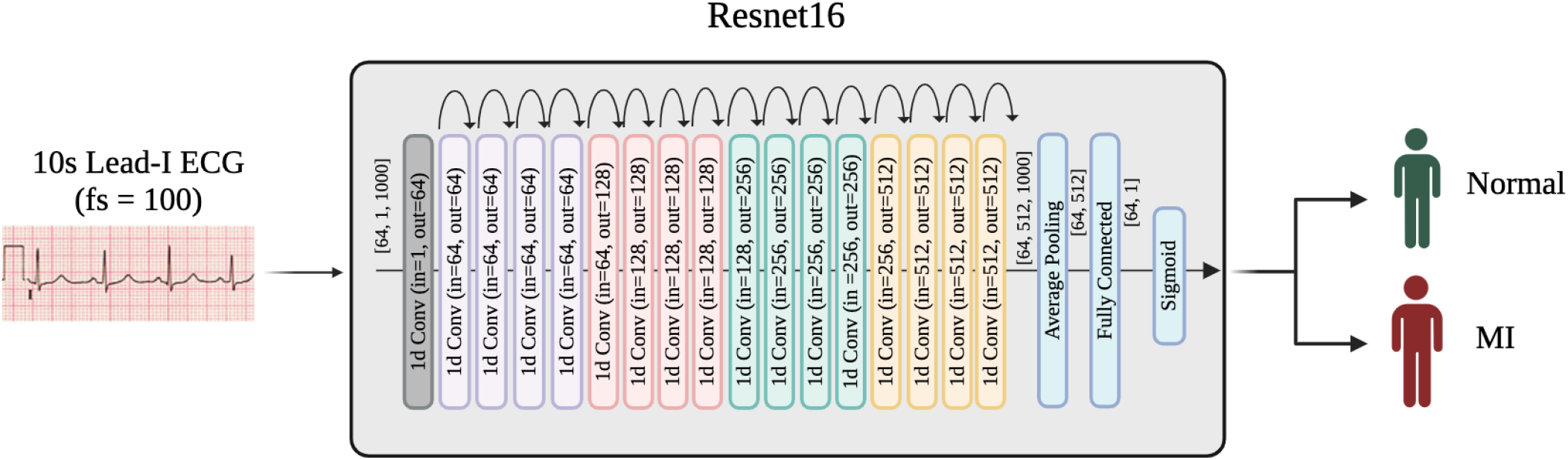
Architecture of our ResNet16 model. In the model comparison task (section III-A), the positive class (“MI”) refers to acute and/or chronic MI, while in the lead comparison task (section III-B), it only refers to acute MI.

From here, we considered two different sets of train-test splits. For the task of model comparison, we used the PTB-XL dataset’s pre-assigned train-val-test folds (80-10-10) which have equal stratifications of Normal and MI, and we disregarded the chronicity of MIs (the classification task was detection of MI regardless of chronicity). However, for the task of comparing different leads in detecting MI, since detection of acute MI is of far greater clinical significance than chronic MI, we modified the split in a way that ensures we only have acute MIs in our test set, and a mixture of acute and chronic MIs in our validation and train sets. We deleted ECG records in the test set which came from patients that also appeared in the train set. This yielded a training set of 4451 normal and 550 MI records, and a test set consisting of 703 normal and 43 acute MI records, which is what is illustrated in Figure 2

For the task of binary classification, we evaluated the performance of 4 different models for input lead-I. In addition to lead-I, we compared the performance of our ResNet16 model trained on different individual leads as input (for the same binary classification task of MI vs. Normal), and benchmarked them against using all 12 leads and the 6 non-precordial leads (I, II, III, aVF, aVL and aVR) used in commercial portable ECG devices. We calculated the area under the receiver-operating curve (AUC), area under the precision-recall curve (AUPR), sensitivity, specificity, and accuracy.

We used signals sampled at 100 Hz, resulting in time series of length 1000, and pre-processed them with a band-pass filter, with a lower bound of 0.5 Hz, and an upper bound of 40 Hz.

### B. Model

For the task of classification of ECG signals, we used a variation of ResNet18 [19] model specifically curated for 1-D signals like ECGs with approximately 18 million parameters. Unlike classic ResNet [20] models used for images that uses 2-D convolutional layers, this model uses 1-D convolutional layers. The model has 16 residual blocks where each residual block consists of a convolutional layer with kernel filters of size 7, batch normalization layer, ReLU activation, and a max pooling layer in the end to reduce the dimensions. The 16 residual blocks are followed by a classification block consisting of a linear layer for the diagnosis alongside a normalization layer and ReLU activation. We had dropouts (dropout rate 0.5) in every block of the model for regularization. The model was trained from random initialization using the Adam optimizer with an exponential learning rate scheduler (lr = 0.001, gamma = 0.97) which ensures convergence by reducing the learning rate at regular intervals. We used weighted binary crossentropy loss with a weight parameter of 5 to account for the class imbalance by increasing the probability of prediction of the non-dominant class.

For comparison, we also employed three other models.

1. ECGTransform: an ECG arrhythmia classification network with bidirectional transformer presented in [9],
2. Pre-trained ResNet [19]: a 1-D ResNet model initialized with weights pre-trained on PhysioNet for classification of Atrial Fibrillation from ECG signals, and
3. CNN18: an 18-layer 1-D CNN model. Each convolutional layer in CNN18 is followed by batch normalization and a ReLU activation function. The convolutional layers use a kernel size of 3 with padding of 1 to maintain the input size. The number of filters (channels) is 64 for the first 17 layers and 128 for the final layer.

For all four models, we performed hyperparameter tuning over learning rate and the gamma variables using the validation set. In Section III we report the result of the best-performing hyperparameter setting for each model on the test set.

## III. Results

### A. Model Comparison

First, we evaluated the performance of different models on the task of MI detection. Table I summarizes the performance of each model on the test set which consists of 703 Normal and 144 MI ECGs. The thresholds for accuracy, precision, and recall were chosen by a clinical expert. The ROC and PR curves are displayed in Fig. 4. Overall, the best performing model is the ResNet16, yielding AUC=0.92, AUPR=0.82, and recall=0.81. Since in this case recall and AUC are of greater clinical significance, we choose ResNet16 for the detection of acute MI, which we explore next.

**TABLE I.**
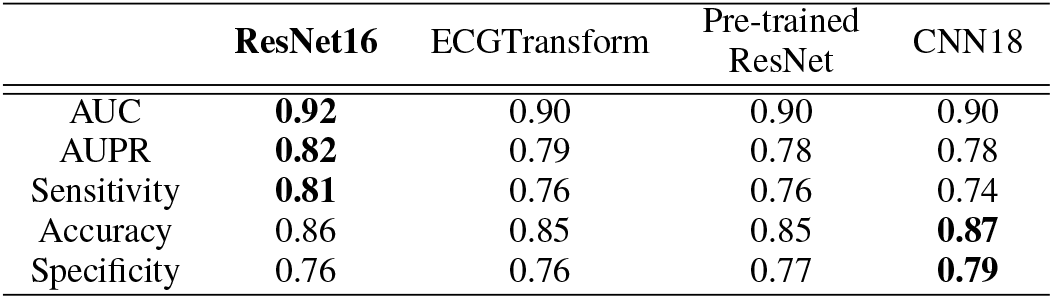
Result comparison of different models

**Fig. 4.**
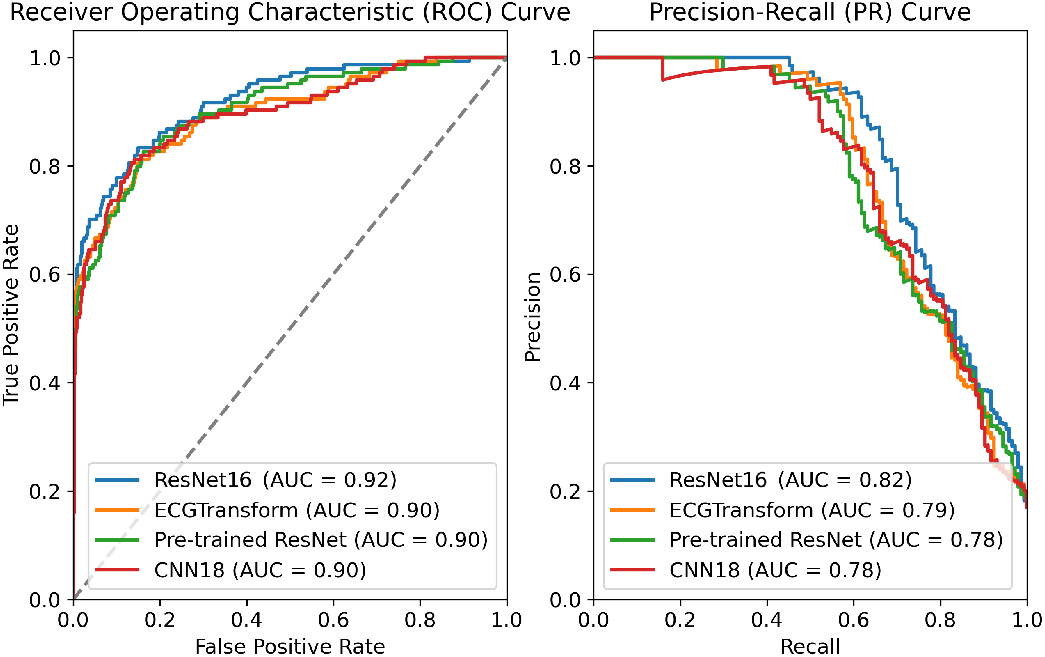
Performance comparison with ROC and PR curves of the models.

### B. Lead Comparison

In order to compare the discriminative power of different leads in the task of detecting acute MI, we trained the ResNet16 model on different input leads, while keeping all hyperparameters the same. The prevelance of MI in our test set for this experiment was 6.8*%*. Table II shows the AUC and AUPR of the test set when the model is trained on each (combination of) input lead. First, and expectedly, the benchmark tasks of training on 6 leads (I, II, III, aVF, aVL and aVR) and all 12 leads achieved a high AUC of 0.99. Among the single-lead predictions, leads V2-V5 performed the best, reflected in their higher AUC, which is in concordance with a previous study [21]. Particularly, lead V2 has the best performance which is consistent with the findings of [14]. We can also see that lead-I has comparably high performance compared to other leads, making it a promising candidate for the task of acute MI detection using wearable devices.

**TABLE II.** Test set performance of the model trained on single leads

| Lead | AUC | AUPR | Sensitivity | Accuracy | Specificity |
| --- | --- | --- | --- | --- | --- |
| I | 0.95 | 0.78 | 0.86 | 0.85 | 0.85 |
| II | 0.91 | 0.69 | 0.84 | 0.89 | 0.89 |
| III | 0.90 | 0.64 | 0.81 | 0.82 | 0.83 |
| aVL | 0.96 | 0.80 | 0.88 | 0.88 | 0.88 |
| aVR | 0.91 | 0.72 | 0.84 | 0.87 | 0.87 |
| aVF | 0.91 | 0.64 | 0.81 | 0.83 | 0.83 |
| V1 | 0.90 | 0.66 | 0.81 | 0.83 | 0.83 |
| V2 | 0.96 | 0.84 | 0.91 | 0.87 | 0.86 |
| V3 | 0.96 | 0.81 | 0.88 | 0.92 | 0.92 |
| V4 | 0.96 | 0.79 | 0.86 | 0.93 | 0.94 |
| V5 | 0.96 | 0.79 | 0.88 | 0.90 | 0.91 |
| V6 | 0.95 | 0.74 | 0.88 | 0.90 | 0.90 |
| 6 leads | 0.99 | 0.89 | 0.91 | 0.97 | 0.98 |
| 12 leads | 0.99 | 0.94 | 0.95 | 0.98 | 0.98 |

## IV. Discussion

While physicians most commonly rely on all 12 leads of the ECG data to detect MI, in this work we demonstrated that lead-I ECG alone can be effectively utilized to detect not only chronic MI but also acute MI (AUC=0.95), which is a more clinically relevant task. This finding is particularly remarkable when considering the relationship between the location of the MI and the leads best suited to detect it. Specifically, MIs are categorized based on the affected region of the heart into four main types: anterior, inferior, lateral, and posterior. In the clinical settings, while lead-I is often used to characterize MI in the lateral region of the heart, it is believed to have limited utility in identifying non-lateral (anterior, inferior, or posterior) MIs. In this dataset, we had less than a handful of MIs that occurred exclusively in the lateral region of the heart, meaning that the grand majority of the MIs were non-lateral. Therefore, our success in detecting mostly non-lateral MIs using only lead-I is a clinically significant and rather unexpected finding. Furthermore, given the significant imbalance of outcomes in our training data (6.8*%* MI), we speculate that the model would perform even better when trained on a more balanced dataset. We believe that our proof-of-concept work holds great potential for clinical deployment. Model inference is quite fast (*<*1s) for each new example, making this suitable for use in clinical settings. Future developments that bring our work closer to clinical deployment would entail not only external validation on other datasets, but also on ECG signals that are acquired directly from wearable devices. Since wearable device ECG signals inherently suffer from higher noise levels, we expect a performance degradation when evaluating the model on those signals. However, conditioned on having sufficiently large wearable device ECG data for fine-tuning the model, we expect to have comparable results to what we presented here. Upon external validation, integrating our algorithm into wearable devices could save millions of lives by prompting timely medical attention.

## Data Availability

All data used are available online at https://physionet.org/content/ptb-xl/1.0.3/

## References

[1] World Health Organization, “Top 10 causes of death,” 2020.

[2] G. De Luca et al., “Time delay to treatment and mortality in primary angioplasty for acute myocardial infarction: every minute of delay counts,” Circulation, vol. 109, no. 10, pp. 1223–1225, 2004.

[3] Z. Gul and A. N. Makaryus, “Silent myocardial ischemia,” 2019.

[4] W. T. Qureshi, Z.-M. Zhang, P. P. Chang, W. D. Rosamond, D. W. Kitzman, L. E. Wagenknecht, and E. Z. Soliman, “Silent myocardial infarction and long-term risk of heart failure: the aric study,” Journal of the American College of Cardiology, vol. 71, no. 1, pp. 1–8, 2018.

[5] E. M. Antman et al., “ACC/AHA guidelines for the management of patients with ST-elevation myocardial infarction: A report of the American College of Cardiology/American Heart Association Task Force on Practice Guidelines (committee to revise the 1999 guidelines for the management of patients with acute myocardial infarction),” Journal of the American College of Cardiology (JACC), vol. 44, no. 3, 2004.

[6] S. Nazarian et al., “Diagnostic accuracy of smartwatches for the detection of cardiac arrhythmia: systematic review and meta-analysis,” JMIR, vol. 23, no. 8, 2021.

[7] N. Isakadze and S. S. Martin, “How useful is the smartwatch ecg?,” Trends in Cardiovascular Medicine, vol. 30, no. 7, pp. 442–448, 2020.

[8] R. Alam et al., “QTNet: Deep learning for estimating QT intervals using a single lead ECG,” in 45th Int Conf of EMBS (EMBC), IEEE, 2023.

[9] H. El-Ghaish and E. Eldele, “ECGTransForm: Empowering adaptive ECG arrhythmia classification framework with bidirectional transformer,” Biomedical Signal Processing and Control, vol. 89, 2024.

[10] R. Alam et al., “Detecting QT prolongation from a single-lead ECG with deep learning,” PLOS Digital Health, vol. 3, no. 6, 2024.

[11] Z. Lin et al., “Automated detection of myocardial infarction using robust features extracted from 12-lead ECG,” Signal, Image and Video Processing, vol. 14, pp. 857–865, 2020.

[12] M.-L. Løchen et al., “Can single-lead computerized electrocardiography predict myocardial infarction in young and middle-aged men? the Tromsø study,” Jour of Cardiovascular Risk, vol. 6, no. 4, 1999.

[13] W.-C. Liu et al., “A deep learning algorithm for detecting acute myocardial infarction: Deep learning model to detect AMI,” EuroIntervention, vol. 17, no. 9, 2021.

[14] C. M. Gibson et al., “Evolution of single-lead ECG for STEMI detection using a deep learning approach,” International Journal of Cardiology, vol. 346, pp. 47–52, 2022.

[15] V. Jahmunah et al., “Automated detection of coronary artery disease, myocardial infarction and congestive heart failure using GaborCNN model with ECG signals,” Comp in Bio and Med, vol. 134, 2021.

[16] A. Alghamdi et al., “Detection of myocardial infarction based on novel deep transfer learning methods for urban healthcare in smart cities,” Multimedia Tools and Applications, pp. 1–22, 2020.

[17] M. Hammad et al., “Myocardial infarction detection based on deep neural network on imbalanced data,” Multimedia Systems, 2022.

[18] P. Wagner, N. Strodthoff, R. Bousseljot, W. Samek, and T. Schaeffter, “PTB-XL, a large publicly available electrocardiography dataset (version 1.0. 1), physionet (2020).”

[19] S. Hong et al., “HOLMES: Health online model ensemble serving for deep learning models in intensive care units,” in Proc. of 26th ACM SIGKDD Int Conf on Knowledge Discovery & Data Mining, 2020.

[20] K. He, X. Zhang, S. Ren, and J. Sun, “Deep residual learning for image recognition,” IEEE CVPR, pp. 770–778, 2016.

[21] A. Gupta et al., “Deep learning for cardiologist-level myocardial infarction detection in electrocardiograms,” 8th Euro Med and Bio Eng Conf (EMBEC), pp. 341–355, 2021.

